# Immunoprecipitation-targeted proteomics assays facilitate rational development of SARS-CoV-2 serological diagnostics

**DOI:** 10.1101/2021.10.25.21265408

**Authors:** Zhiqiang Fu, Yasmine Rais, Andrei P. Drabovich

## Abstract

Current design of serological tests employs conservative immunoassay approaches and is often focused on convenience, speed of manufacturing, and affordability. Limitations of such serological tests include semi-quantitative measurements, lack of standardization, potential cross-reactivity, and inability to distinguish between antibody subclasses. As a result of cross-reactivity, diagnostic specificity of serological antibody tests may not be sufficiently high to enable screening of the general asymptomatic populations for the acquired immunity against low-prevalence infectious diseases, such as COVID-19. Likewise, lack of a single standard for assay calibration limits inter-laboratory and international standardization of serological tests. In this study, we hypothesize that combination of immunoaffinity enrichments with targeted mass spectrometry measurements would enable rational design of serology diagnostics of infectious diseases, such as COVID-19. The same instrumental platform allows for sensitive and specific measurements of viral protein antigens, as wells as anti-viral antibodies circulating in human serum. Our proof-of-concept immunoprecipitation - parallel reaction monitoring (IP-PRM) assays quantified NCAP_SARS2 protein with a limit of detection of 313 pg/mL in serum. In addition, a multiplex IP-selected reaction monitoring (IP-SRM) assay facilitated differential quantification of anti-SARS-CoV-2 antibody isotypes and subclasses in patient sera. Simultaneous evaluation of numerous antigen-antibody subclass combinations revealed a receptor-binding domain (RBD)-IgG1 as a combination with the highest diagnostic specificity and sensitivity. Anti-RBD IgG1, IgG3, IgM and IgA1 subclasses, but not IgG2, IgG4 and IgA2, were found elevated in COVID-19-positive sera. Synthetic heavy isotope-labeled peptide internal standards as calibrators revealed elevated anti-RBD IgG1 in positive (510-6700 ng/mL; 0.02-0.22% of total serum IgG1) versus negative sera (60 [interquartile range 41-81] ng/mL). Likewise, anti-RBD IgM was elevated in positive (190-510 ng/mL; 0.06-0.16% of total serum IgM) versus negative sera (76 [31-108] ng/mL). Further validation of immunoprecipitation-targeted proteomics assays as a platform for serological assays will facilitate standardization and improvement of the existing serological tests, enable rational design of novel tests, and offer tools for comprehensive investigation of antibody isotype and subclass cooperation in immunity response.

## INTRODUCTION

Conventional diagnostics of viral infections, such as COVID-19, relies on detection of viral genomes by the polymerase chain reaction (PCR) or the reverse transcription polymerase chain reaction (RT-PCR). Limitations of RNA measurements by RT-PCR include RNA degradation in biological fluids^1^, relatively high false negative rates^2^, and lack of prognostic information^3^. Alternative assays for diagnosis and prognosis of viral infections include serological assays which rely on detection of protein antigens or anti-viral antibodies in biological fluids or blood serum. The most common serological assays measure anti-viral immunoglobulins in blood and enable detection of past and chronic infections, provide prognostic information, and evaluate patient immune status.^4,5^ There is also an increasing number of serological assays measuring circulating viral proteins, to enable screening of donor blood for chronic infections or differential diagnosis of related viruses^6^. Recently, combined serological test for simultaneous detection of circulating protein antigens and anti-viral antibodies were developed to complement RT-PCR diagnostics, or facilitate earlier detection of viral infections.^7-9^

Enzyme linked immunosorbent assay (ELISA) and lateral flow immunoassay (LFA), the most common tools for serology diagnostics, present highly sensitivity, robust and convenient analytical assays to measure viral proteins and anti-viral immunoglobulins in clinical samples^10,11^. Immunoassay limitations, however, include non-specific binding, insufficient diagnostics specificity, lack of high-quality international reference standards, challenges with multiplexing, and antibody cross-reactivity to the highly homologous proteins of related strains^12^. Differential measurement of the full set of immunoglobulin isotypes (total IgG, total IgA, IgM, IgE, IgD) and subclasses (IgG1, IgG2, IgG3, IgG4, IgA1 and IgA2) by indirect ELISA requires eleven individual measurements for each patient sample and is not routinely performed. As a result, common serological tests do not evaluate the isotype- and subclass-specific humoral immune response which could provide additional diagnostic and prognostic information. It has been well established that the identity and circulating levels of immunoglobulin isotypes and subclasses depend on numerous factors, such as time after exposure (early response IgM), antigen identity (peptides or polysaccharides), route of infection (respiratory, urinary, or topical), cell-mediated immunity (class switching induced by either type 1 or 2 helper T cells), subclass stability (reduced half-life of IgG3), and different effector functions (cytotoxicity or phagocytosis)^13,14^.

Mass spectrometry (MS) with its near-absolute analytical selectivity and multiplexing capabilities presents an alternative approach for serology diagnostics. MS has recently been used for identification and quantification of SARS-CoV-2 proteins in biological and clinical samples^15-18^. Without extensive fractionation, however, MS assays presented relatively poor analytical sensitivity and resulted in low diagnostic sensitivity.

In this work, we hypothesized that combination of immunoaffinity enrichments and mass spectrometry measurements could resolve common limitations of immunoassays and mass spectrometry assays. We suggested that combined immunoprecipitation-selected reaction monitoring (IP-SRM) or immunoprecipitation-parallel reaction monitoring (IP-PRM) assays could facilitate sensitive and selective quantification of SARS-CoV-2 protein antigens and anti-SARS-CoV-2 antibodies in patient samples. Proposed assays (**Figure 1)** may provide a single platform for: (i) quantification of SARS-CoV-2 proteins in patient samples, to complement RT-PCR diagnostics and develop prognostic tests; (ii) differential quantification of SARS-CoV-2 emerging mutants; (iii) differential quantification of anti-SARS-CoV-2 antibody isotypes (IgG, IgA, IgM) and subclasses (IgG1-4, IgA1-2); (iv) rational design of serological diagnostics through selection of antigen-antibody subclass combinations with the highest diagnostic specificity and sensitivity; and (v) standardisation of SARS-CoV-2 protein and antibody assays via stable, pure, and affordable synthetic peptide internal standards.

**Figure 1.**
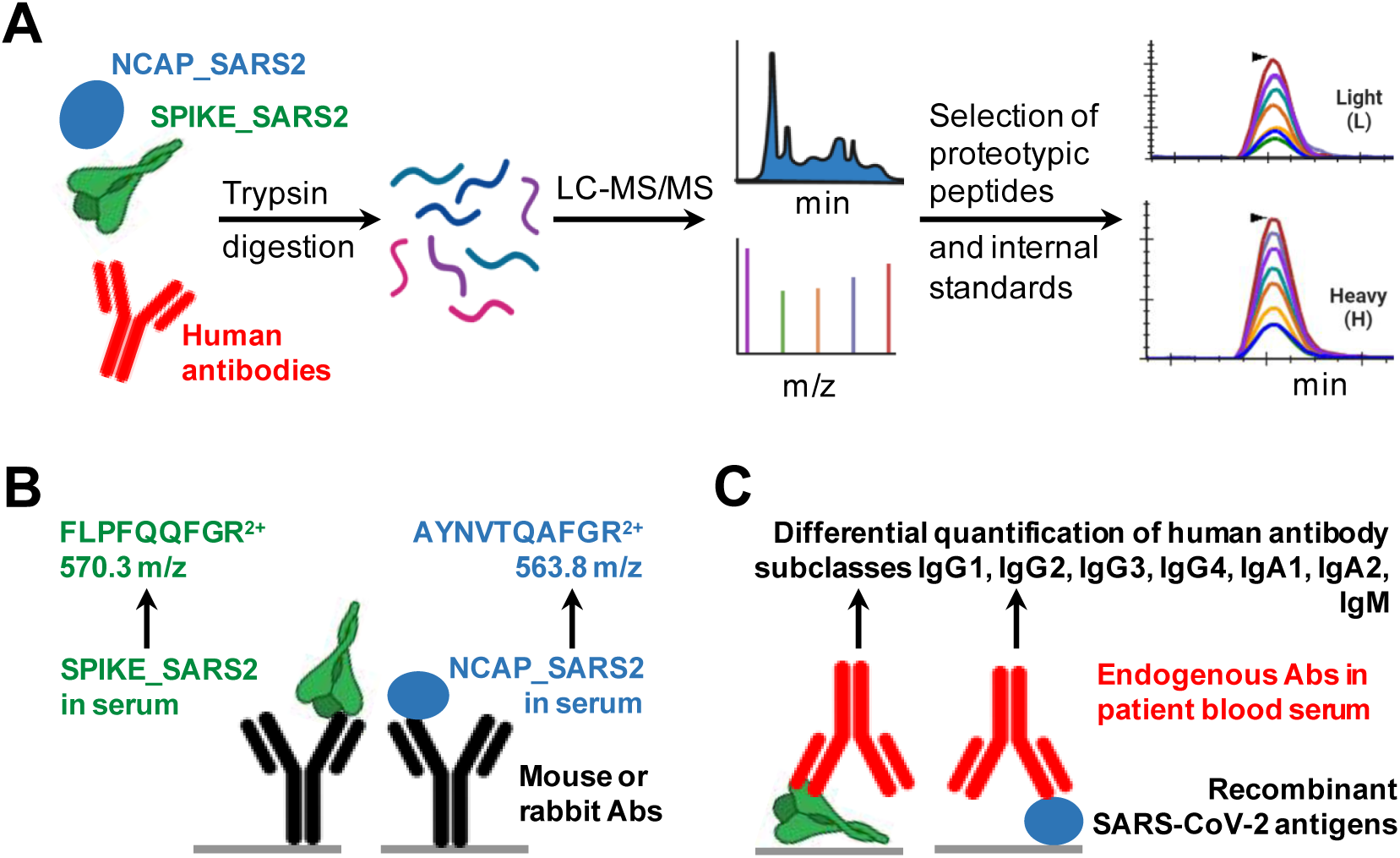
Design of immunoaffinity-mass spectrometry assays for serological diagnostics. **(A)** Identification of proteotypic peptides by shotgun LC-MS/MS and development of quantitative targeted proteomics assays. Recombinant SPIKE_SARS2 and NCAP_SARS2 proteins, or human antibodies, are digested by trypsin, tryptic peptides are analyzed by liquid chromatography-shotgun mass spectrometry (LC-MS/MS), the best proteotypic peptides are identified and synthesized as heavy isotope-labeled internal standards, and SRM transitions with the highest signal-to-noise ratio are selected. **(B)** Setup of IP-SRM or IP-PRM assays for quantification of SPIKE_SARS2 and NCAP_SARS2 proteins. (**C**) Setup of IP-SRM assays for quantification of anti-SARS-CoV-2 immunoglobulin individual subclasses (IgG1, IgG2, IgG3, IgG4, IgA1, IgA2, and IgM).

## RESULTS

### Development of IP-PRM assays for quantification of SARS-CoV-2 proteins

To develop PRM assays for SARS-CoV-2 proteins, we obtained recombinant proteins, identified tryptic peptides by shotgun mass spectrometry^21^, prioritized the most intense peptides based on the label-free quantification with MaxQuant, and selected the most intense transitions with Skyline. In addition, we re-searched several publically available SARS-CoV-2 proteomic datasets^22,23^, confirmed the choice of proteotypic peptides, and applied label-free iBAQ quantification to determine relative abundances of SARS-CoV-2 proteins: NCAP_SARS2 (nucleoprotein; 55% of the viral proteome), VME1_SARS2 (membrane protein; 18%), AP3A_SARS2 (ORF3a protein; 9%), SPIKE_SARS2 (spike glycoprotein; 8%), ORF9B_SARS2 (ORF9b protein; 7%), NS7A_SARS2 (ORF7a protein; 1.2%), NS6_SARS2 (ORF6 protein; 0.7%), NS8_SARS2 (ORF8 protein; 0.4%), and other proteins (∼0.4%). A combined database of SARS-CoV-2 proteins, tryptic peptides, and MS fragmentation spectra facilitated rapid development of the targeted proteomics assays. Best proteotypic peptides (**Figure 1B**) were synthesized as heavy isotope-labeled peptides and used as internal standards for development of PRM assays and quantification of SPIKE_SARS2 and NCAP_SARS2. Selection of AYNVTQAFGR (NCAP_SARS2) and FLPFQQFGR (SPIKE_SARS2) as the best proteotypic peptides confirmed previous studies^22^. To develop IP assays, we tested three different anti-NCAP_SARS2 and two anti-SPIKE_SARS2 antibodies, and evaluated assay performance with the corresponding recombinant proteins spiked into human serum (**Figure 2**). As a result, our IP-SRM assays detected 1.25 ng/mL of SPIKE_SARS2 (238 amol on column) and 313 pg/mL of NCAP_SARS2 (170 amol on column) in serum (**Figure 3**). These levels were comparable to a recent study which identified AYNVTQAFGR as the best proteotypic peptide of NCAP_SARS2, and reported LOD of 200 amol on column for quantification of NCAP_SARS2 protein in nasopharyngeal swab samples^23^. A recent study reported that the median levels of NCAP_SARS2 protein in capillary blood were 3,896 pg/mL for pre-symptomatic and 1,931 pg/mL for symptomatic patients^24^, well above the limit of detection of our IP-PRM assay.

**Figure 2.**
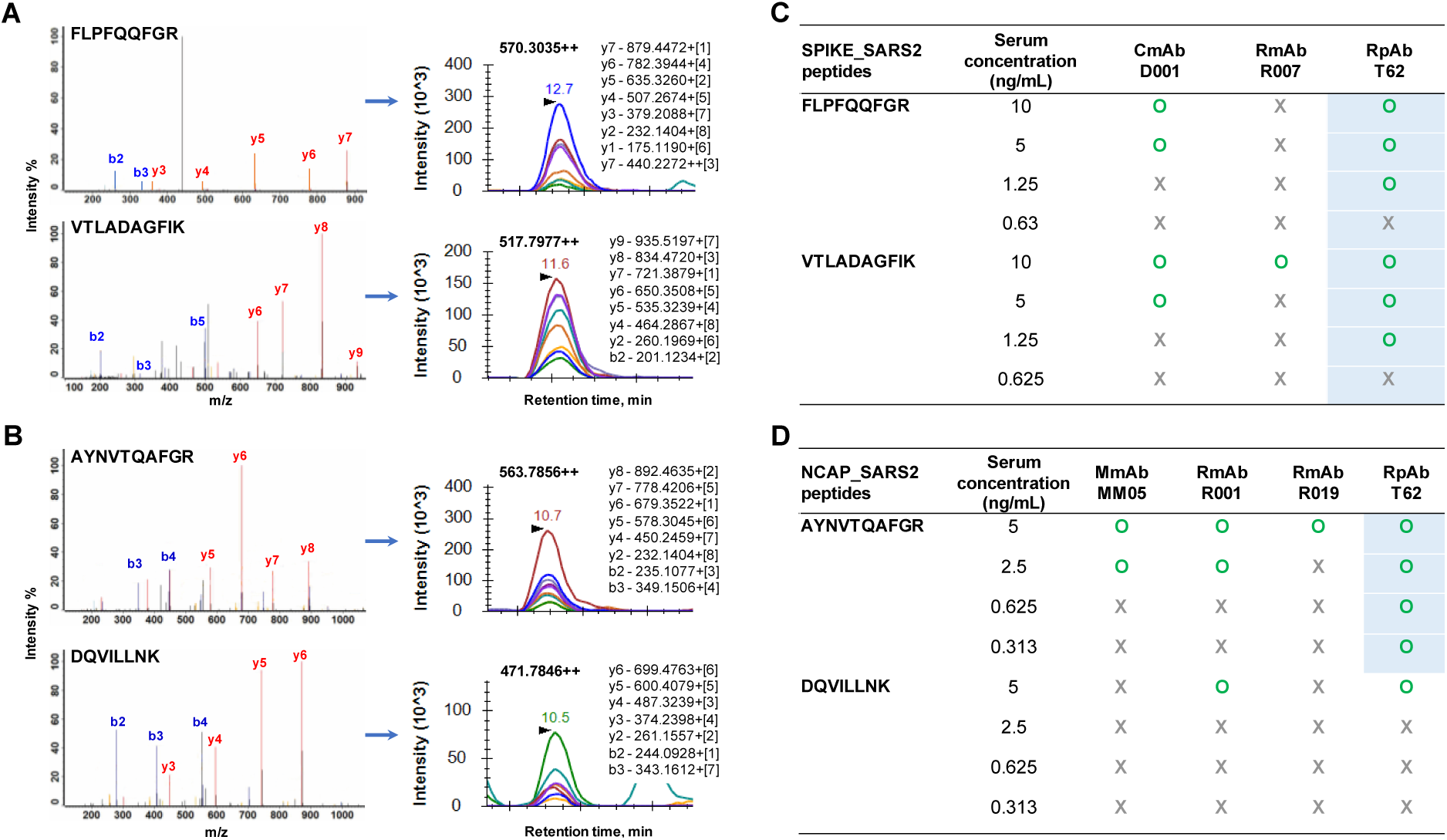
Development of IP-PRM assays for quantification of SPIKE_SARS2 and NCAP_SARS2 proteins. Selection of proteotypic peptides and PRM transitions for quantification of SPIKE_SARS2 (**A**) and NCAP_SARS2 (**B**) proteins. (**C**) SPIKE_SARS2 immunoprecipitation-PRM assays developed with anti-SPIKE_SARS2 chimeric monoclonal antibody CmAb (D001), rabbit monoclonal RmAb (R007) and rabbit polyclonal antibody RpAb (T62). (**D**) NCAP_SARS2 immunoprecipitation-PRM assays developed with anti-NCAP_SARS2 mouse monoclonal antibody MmAb (MM05), rabbit monoclonal antibodies RmAb (R001) and RmAb (R019), and rabbit polyclonal antibody RpAb (T62).

**Figure 3.**
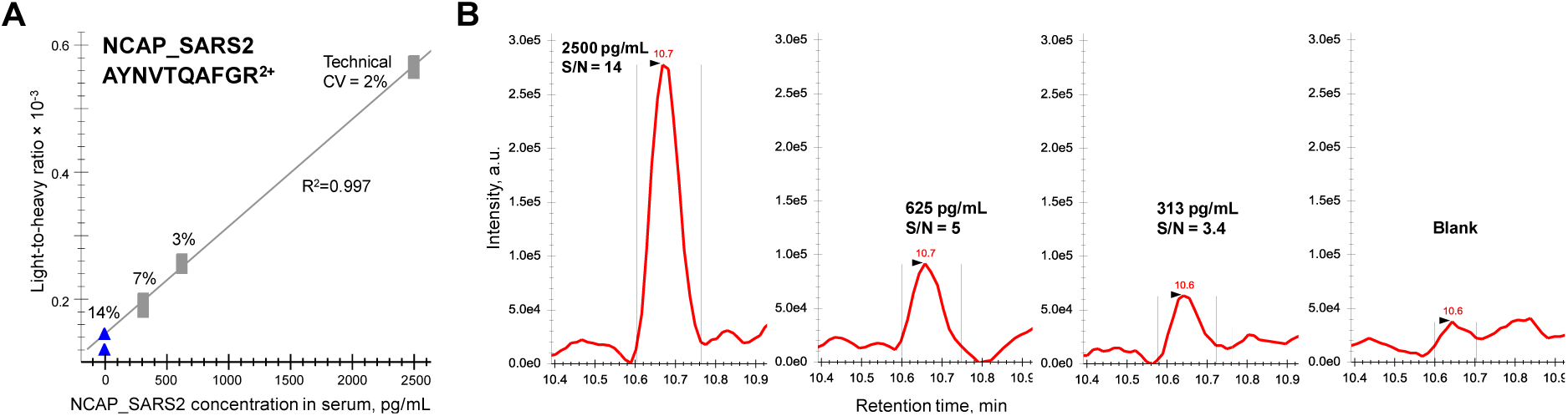
Quantification of recombinant NCAP_SARS2 protein spiked into human serum. Immunoprecipitation-PRM assay with a rabbit polyclonal antibody RpAb T62 revealed a linear response **(A)** and a limit of detection of 313 pg/mL (S/N>3) in serum, or 170 amol on column (**B**). A recent study reported that the median levels of NCAP_SARS2 protein in capillary blood were 3,896 pg/mL for pre-symptomatic and 1,931 pg/mL for symptomatic patients^24^, and well above the limit of detection of our IP-PRM assay.

### Quantification of viral proteins in serum by ELISA

To develop an in-house ELISA, we tested the performance of three anti-SPIKE_SARS2 and four anti-NCAP_SARS2 antibodies. We first evaluated all combinations of these antibodies to capture (200 ng/well) and detect (20 ng/well; biotinylated in-house) corresponding proteins (**Figure 4**). Antibody pairs which provided the highest signal intensity (OD at 450 nm) for the recombinant SPIKE_SARS2 (monomeric S1+S2 ECD) and NCAP_SARS2 diluted in the sample buffer were then re-evaluated with proteins spiked into human serum. The best pairs of antibodies included: (i) a capture mouse/human chimeric monoclonal antibody (CmAb D001) and a detection rabbit polyclonal antibody (RpAb T62) to measure SPIKE_SARS2 with LoQ of 63 pg/mL (66 amol/well) in human serum, and (ii) a capture rabbit polyclonal antibody (RpAb T62) and a detection mouse monoclonal antibody (MmAb MM05) to measure NCAP_SARS2 with LoQ of 31 pg/mL (47 amol/well) in human serum (**Figure 4**). Some commercial ELISA kits (for example, Sino Biological) would have comparable sensitivity (7.8 pg/mL for SPIKE_SARS2 RBD corresponding to 40 pg/mL for S1+S2 ECD).

**Figure 4.**
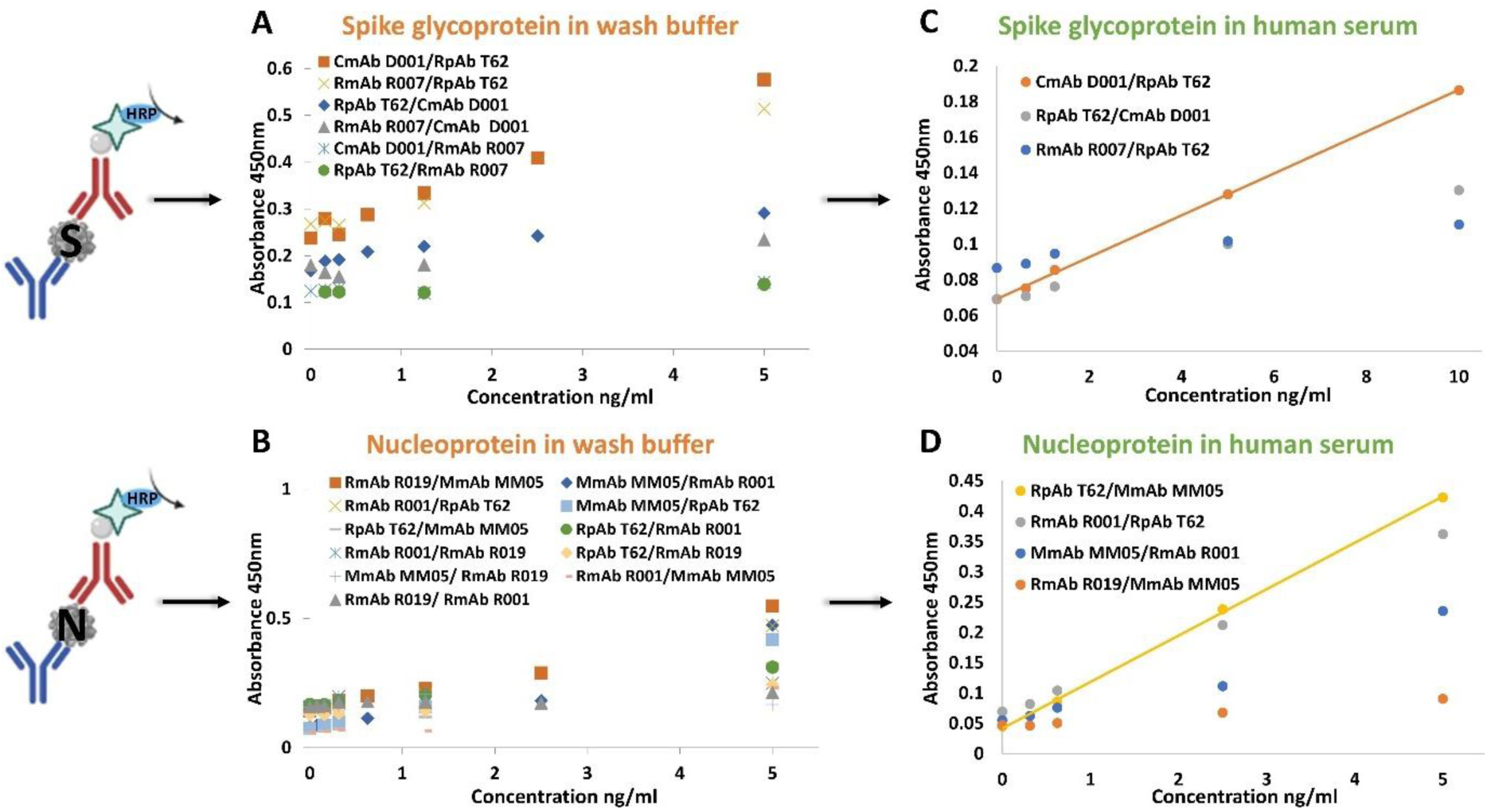
Development of in-house ELISA for measurement of SPIKE_SARS2 and NCAP_SARS2 proteins in human serum. Numerous combinations of capture and detection anti-SPIKE_SARS2 (**A**) and anti-NCAP_SARS2 (**B**) antibodies were evaluated. The best pairs of antibodies included a capture mouse/human chimeric monoclonal antibody (CmAb D001) and a detection rabbit polyclonal antibody (RpAb T62) to measure SPIKE_SARS2 with LoQ of 63 pg/mL in serum (**C**), and a capture rabbit polyclonal antibody (RpAb T62) and a detection mouse monoclonal antibody (MmAb MM05) to measure NCAP_SARS2 with LoQ of 31 pg/mL in serum (**D**).

### Development of IP-SRM assays for the differential quantification of anti-SARS-CoV-2 immunoglobulins

Our approach for the differential quantification of human immunoglobulin isotypes (IgG, IgM, and IgA) and subclasses (IgG1, IgG2, IgG3, IgG4, IgA1 and IgA2) relied on measurements of the unique proteotypic peptides within the constant heavy chains (**Supplemental Figure S1**). To select unique proteotypic peptides for each isotype and subclass, we searched our previous proteomic datasets^25^, Peptide Atlas data^26^ and literature data^27,28^. Selected proteotypic peptides represented all immunoglobulin allotypes^14^. Absence of high-frequency polymorphic missense variants was confirmed with the GnomAD database v2.1.1^29^ (**Table S4**). The search of NextProt database^30^ ensured absence of glycosylation sites or other post-translational modifications within the sequences of proteotypic peptides. Finally, nine synthetic heavy isotope-labeled peptides were used for SRM assay development (**Table S4**). SRM assays for quantification of IGHG1 and IGHM in serum revealed LoQs of 0.3 *f*mol on column for IGHG1 peptide (corresponding to 14 ng/mL of IgG1) and 1 *f*mol on column for IGHM peptide (corresponding to 62 ng/mL of IgM) (**Figure 5 and Table S7**).

**Figure 5.**
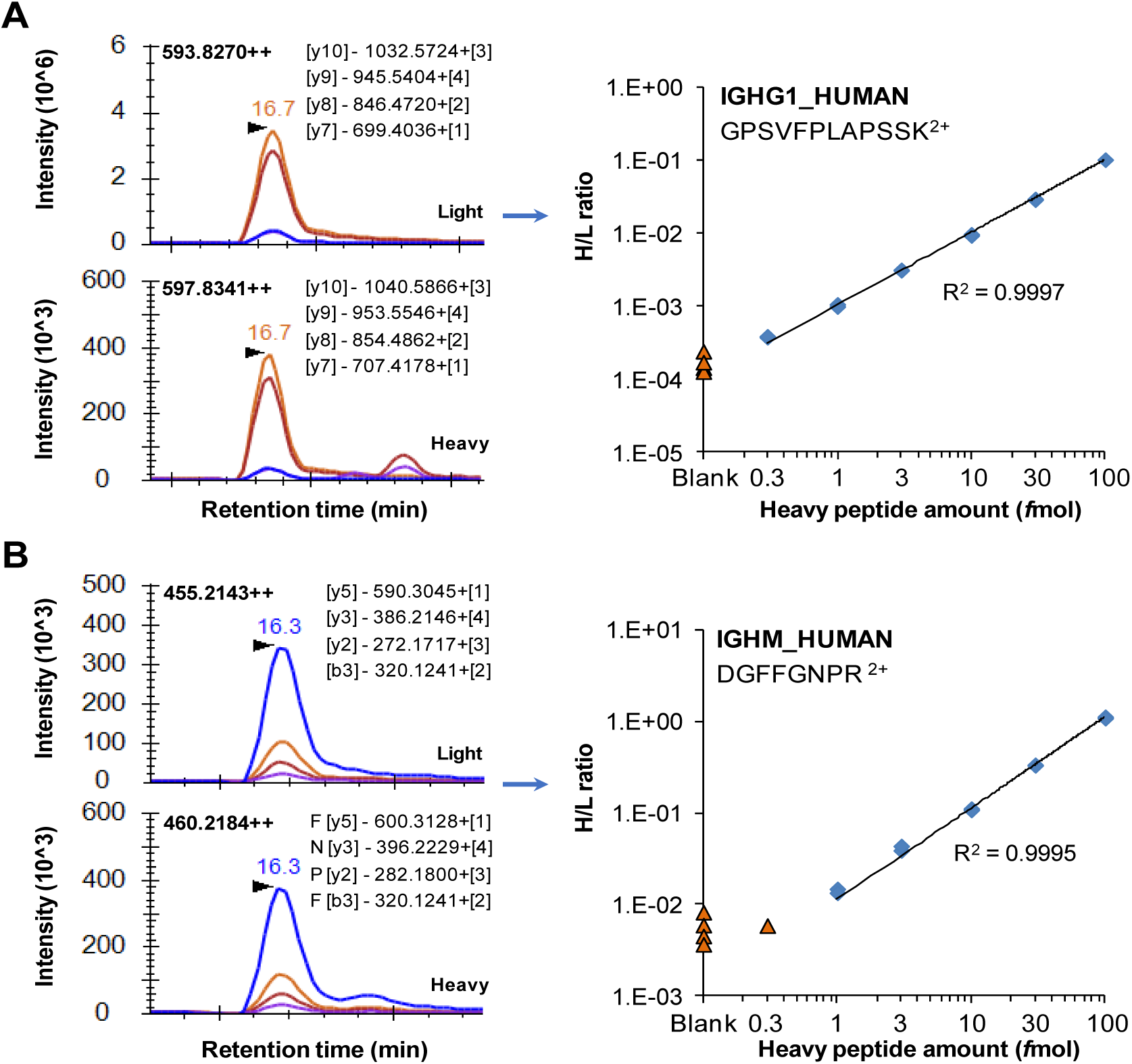
Representative SRM assays for quantification of IGHG1_HUMAN (A) and IGHM_HUMAN (B) in human serum. Unique tryptic peptides of a CH1 region of heavy constant gamma 1 and a CH2 region of heavy constant mu represented total IgG1 and IgM, respectively. Calibration curves corresponded to dilution series of heavy isotope labeled peptide internal standards spiked into digest of human serum and revealed limits of detection of 0.3 and 1 *f*mol on column for IgG1 and IgM, respectively.

### Quantification of COVID-19-specific immunoglobulins by IP-SRM

COVID-19-specific immunoglobulins (IgG1, IgG2, IgG3, IgG4, IgM, IgA1, and IgA2) were quantified with NCAP_SARS2 and SPIKE_SARS2 S1+S2 ECD, S1 and RBD proteins in 7 negative and 3 positive sera (**Table S8**).

### Quantification of COVID-19-specific immunoglobulins by ELISA

We developed an in-house indirect ELISA and measured relative abundance of SARS-CoV-2-specific IgG and IgGAM immunoglobulins captured by NCAP_SARS2 and SPIKE_SARS2 S1+S2 ECD, S1 and RBD proteins. Both IgG or IgGAM assays were sensitive and specific enough to differentiate between 7 negative and 3 positive sera (**Figures 6B** and **S2**).

**Figure 6.**
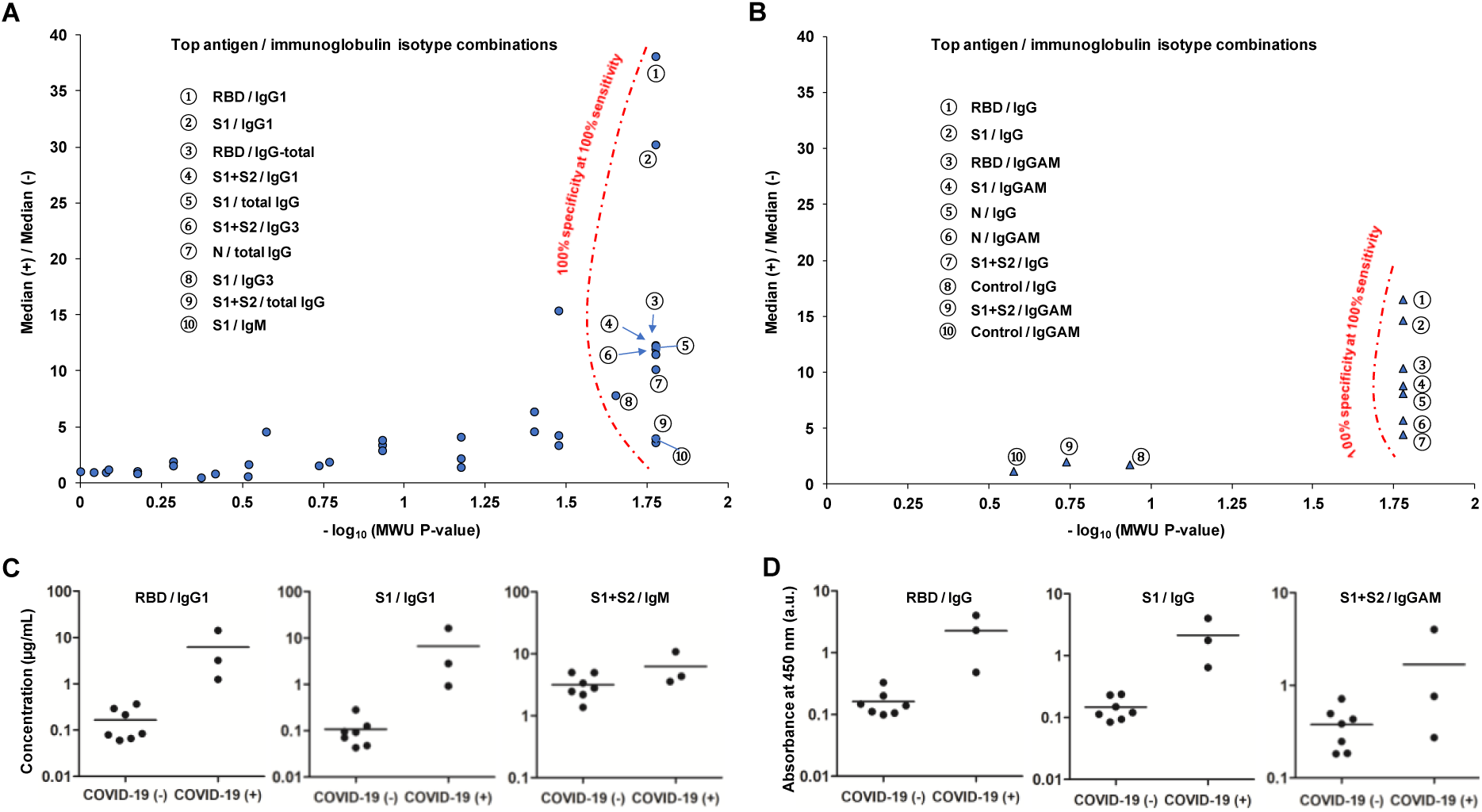
Rational design of SARS-CoV-2 serological tests. Roadmaps for evaluation of numerous antigen-immunoglobulin isotype/subclass combinations and selection of pairs which provided 100% diagnostic specificity at 100% diagnostic sensitivity based on measurements by IP-SRM (**A**) and indirect ELISA (**B**). For the COVID-19 positive versus negative serum samples, RBD/IgG1 and S1/IgG1 combinations measured by IP-SRM provided the highest median fold differences and dynamic range (e.g. higher true-positive signal and lower background). Evaluated immunoglobulin subclasses and isotypes included IgG1, IgG2, IgG3, IgG4, IgM, IgA1, IgA2, total IgG, and total IgA for IP-SRM and, IgG and IgGAM for indirect ELISA. Evaluated antigens included recombinant NCAP_SARS2 (N) and recombinant SPIKE_SARS2 receptor-binding domain sequence (RBD), S1 subunit sequence (S1), and an extracellular domain sequence (S1+S2). Representative antigen/immunoglobulin isotype combinations with excellent (RBD/IgG1, S1/IgG1) and poor (S1+S2/IgM) diagnostic performance of serology assays based on measurements by IP-SRM (**C**) and indirect ELISA (**D**).

### Selection of antigen-antibody combinations with the highest diagnostic performance

We evaluated numerous antigen - immunoglobulin subclass combinations by IP-SRM and indirect ELISA (**Figure 6)**. Simultaneous evaluation of 36 combinations was a definite advantage of a multiplex IP-SRM assay, and allowed identifying 10 combinations which satisfied two essential criteria: statistically-significant difference (MWU *P-*value<0.05) and no overlap between groups (100% diagnostic specificity at 100% diagnostic sensitivity). Additional ranking by the ratio of medians facilitated selection of combinations with the highest signal-to-noise ratio. RBD/IgG1 and S1/IgG1 were identified as top combinations. Indirect ELISA with anti-IgG and anti-IgGAM detection antibodies confirmed the best performance of RBD/IgG and S1/IgG combinations (**Figure 6B** and **Tables S9**), which was in agreement with the previous studies^31^. It should be noted that S1+S2 ECD antigen revealed poor diagnostic performance (**Figure 6C, D**) due to the high background in negative samples. In addition to the rapid evaluation of numerous combinations, we revealed that IP-SRM assay provided a 2.3-fold wider dynamic range in comparison to ELISA. An IP step of our assay provides a dynamic range of detection ∼2-3 orders of magnitude, similar to typical affinity interactions^32^.

In ELISA, this dynamic range could be further reduced by equilibration parameters and non-specific interactions of the anti-IgG or anti-IgGAM secondary detection antibodies. Unlike immunoassays, SRM measurements (dynamic range of 5-6 orders) allow for the direct quantification of immunoglobulin heavy chains with no cross-reactivity (lower background) and independent of the secondary antibody (no signal loss due to dissociation), thus providing a wider dynamic range. A wider dynamic range of serological diagnostics by IP-SRM could facilitate earlier detection of the triggered immune response and more accurate measurements of its temporal dynamics.

### COVID-19-specific versus total immunoglobulin profiles

Total immunoglobulins (IgG1, IgG2, IgG3, IgG4, IgM, IgA1 and IgA2) were quantified by SRM in 7 negative and 3 positive sera. Median concentrations of total IgG1 and IgM in serum, 2.8 and 0.24 mg/mL, respectively (**Figure 7**) were in agreement with the previously reported ranges for IgG1 (2.8 - 8.2 mg/mL) and IgM (0.2∼2.3 mg/mL)^33-35^. Anti-RBD IgG1, IgG3, IgM and IgA1 subclasses, but not IgG2, IgG4 and IgA2, were found elevated in positive convalescent sera (**Figure 7** and **Tables S8**). Anti-RBD IgG1 was elevated in positive (510-6700 ng/mL; 0.02-0.22% of total serum IgG1) versus negative sera (60 [interquartile range, IQR, 41-81] ng/mL). Anti-RBD IgG1 levels measured by our IP-SRM well correlated (R^2^=0.98) with IgG levels independently measured by SARS-CoV-2 IgG seroconversion ELISA (Innovative Research). Anti-RBD IgM was elevated in positive (190-510 ng/mL; 0.06-0.16% of total serum IgM) versus negative sera (76 [IQR 31-108] ng/mL). Anti-RBD IgG3 was elevated in positive (29-220 ng/mL; 0.004-0.049% of total IgG3) versus negative sera (22 [IQR 13-26] ng/mL). Some positive serum samples revealed anti-RBD IgA1 (35-260 ng/mL; 0.003-0.028% of total IgA1).

**Figure 7.**
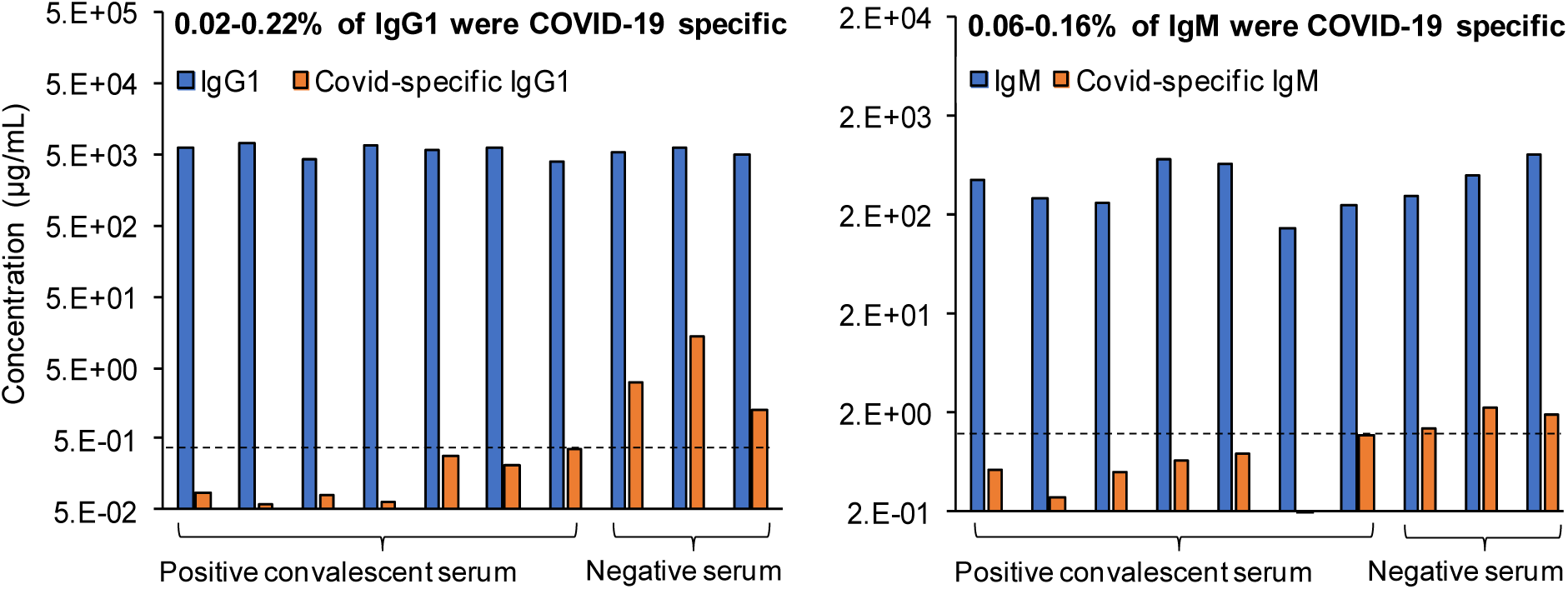
Measurement of total IgG1 and IgM by SRM, and specific anti-RBD IgG1 and IgM by IP-SRM. Serum samples were obtained from COVID-19 negative patients (collected before December 2019) and COVID-19 positive patients (confirmed by RT-PCR). Median levels of total IgG1 and IgM, as measured by SRM in serum samples, were 2.8 and 0.24 mg/mL, respectively. Anti-RBD IgG1 levels were elevated in positive convalescent serum (510-6,700 ng/mL; 0.02-0.22% of total serum IgG1) versus negative serum (60 ng/mL; interquartile range 41-81 ng/mL). Anti-RBD IgG1 levels measured by our IP-SRM well correlated (R^2^=0.98) with IgG levels independently measured by SARS-CoV-2 IgG seroconversion ELISA (Innovative Research). Anti-RBD IgM levels were elevated in positive (190-510 ng/mL; 0.06-0.16% of total serum IgM) versus negative serum (76 ng/mL; interquartile range 31-108). Non-specific binding levels of IgG1 (median 56 ng/mL) and IgM (median 63 ng/mL), as measured with PBS buffer instead of RBD antigen, were subtracted.

## DISCUSSION

Infectious disease diagnostics has been revolutionized with the advent of PCR and RT-PCR. Further developments of infectious disease diagnostics are increasingly utilizing protein antigen and antibody measurements to aid nucleic acids tests and provide additional diagnostic and prognostic information. Hepatitis B testing is one of the prominent examples and utilizes serum measurements of viral DNA by PCR and immunoassay measurements of a viral surface antigen protein, two IgG antibodies against surface and core antigens, and IgM antibody against a core antigen protein^9^. Different combinations of positive and negative results provide detailed interpretation of Hepatitis B infection status (acute, chronic, immune due to vaccination, immune due to previous infection, susceptible to infection, resolved infection).

Standard serological assays to measure anti-pathogen antibodies in blood serum or plasma have been unchanged for decades. The design of serological assays was determined by conservative immunoassay approaches and was often focused on convenience, speed of manufacturing, and affordability. Limitations of such assays included semi-quantitative measurements due to the lack of “gold” standards, potential cross-reactivity, and inability to distinguish between antibody subclasses. Since reference samples with the standardized amounts of anti-pathogen antibodies or recombinant monoclonal antibodies are typically not available at the early stages of the novel pathogen epidemics, different laboratories calibrate serology tests with different convalescent serum samples obtained from the recovered patients. Lack of a single reference standard for assay calibration limits inter-laboratory and international standardization of serological tests, and is a recognized limitation. As a result of cross-reactivity, diagnostic specificity of serological antibody tests may not be sufficiently high to enable screening of the general asymptomatic populations for the acquired immunity against low-prevalence infectious diseases, such as COVID-19. For instance, 95% diagnostic specificity of COVID-19 serological tests^36,37^ and 0.3% prevalence in early 2020 in Canada would account for 5% positive predictive value. Interestingly, 90% positive predictive value at such low prevalence could only be achieved with a superior test with 99.97% diagnostic specificity.

PCR and RT-PCR are undoubtedly the techniques with unprecedented analytical sensitivity. Sensitivity of protein assays, however, could be leveraged by the presence of numerous analyte copies (for example, ∼1,000 copies of NCAP_SARS2 protein per virion^38^), longer elimination half-life of proteins (for instance, circulating SARS-CoV proteins were detectable in serum on day 25 after infection onset, while serum RNA was undetectable on day 20^39^), and the higher stability during sample preparation. The most abundant SARS-CoV-2 proteins were recently evaluated as diagnostic and prognostic biomarkers^24^, but evaluation of accessory proteins is still pending, mostly due to the lack of high-quality antibodies and immunoassays. Mass spectrometry has previously been used for identification and quantification of viral proteins in clinical samples^15-17^, and will facilitates evaluation of SARS-CoV-2 accessory proteins as biomarkers. Since rational development of protein biomarkers involves numerous stages of verification and validation, mass spectrometry assays, due to their rapid design and execution, are particularly useful at the early phases of biomarker development^40-43^. Without extensive fractionation, however, mass spectrometry assays present relatively poor analytical sensitivity, resulting in low diagnostic sensitivity.

In this study, we hypothesized that combination of immunoaffinity enrichments with mass spectrometry quantification provides a single instrumental platform for sensitive and specific serological testing for pathogen antigen proteins and anti-pathogen antibodies. Immunoaffinity-mass spectrometry platform enables rational design of serological diagnostics of infectious diseases, provides assay standardization, resolves certain limitations of standard PCR and immunoassay techniques, and facilitates independent evaluation of cross-reactivity of serological immunoassays^44^. Proposed IP-SRM assays combine advantages of two worlds: immunoassays with high analytical sensitivity, and SRM assays with their near-absolute analytical specificity^44-49^. We previously demonstrated that SRM and PRM targeted proteomic assays provided robust tools for quantification of proteins in human cell lines^50,51^, primary cells^52-54^, tissues^55^, various biological fluids^46,56-61^, and serum^44,62^. Additional immunoprecipitation provides up to a 1,000-fold gain in sensitivity, reaching 100 pg/mL levels in complex biological and clinical samples^44^. All steps of IP and sample preparation are implemented on 96-well plates and provide sufficient throughput and reproducibility^25,48^. High reproducibility (coefficient of variation, CV<10%) and throughput (∼100 samples per day) of IP-SRM assays enable reliable quantification of proteins in clinical samples.

We have previously developed IP-SRM for novel proteins^44,47,48,55^ and demonstrated limits of quantification as low as 100 pg/mL in serum^44^. Such sensitivity is sufficient to quantify viral antigens and antiviral antibodies in serum. For example, patients with chronic hepatitis B presented serum surface antigen levels of 40,000 ng/mL^63^, while SARS-CoV nucleocapsid protein was detected at 100-3,200 pg/mL in serum of infected patients^64^. Our proof-of-concept IP-PRM assays measured NCAP_SARS2 protein with a limit of detection of 313 pg/mL in serum. Likewise, antiviral IgG immunoglobulins in serum present a range of 1-100 µg/mL^24,65^, and a recent semi-quantitative SARS-CoV-2 IgG serological test has a clinical cutoff of 0.77 μg/mL^66^. Our IP-SRM measurements with synthetic peptide internal standards as calibrators revealed elevated anti-RBD IgG1 (510-6,700 ng/mL), IgG3 (29-220 ng/mL), IgM (190-510 ng/mL), and IgA1 (up to 260 ng/mL in some samples), but not IgG2, IgG4 or IgA2 immunoglobulins. Our multiplex IP-SRM assay facilitated simultaneous evaluation of 36 antigen-antibody subclass combinations, and revealed RBD-IgG1 as a combination with the highest diagnostic specificity and sensitivity. These data were in agreement with our in-house indirect ELISA which confirmed RBD/IgG as the top combination.

It should be mentioned that differential quantification of a full set of immunoglobulin isotypes (IgG, IgA, IgM, IgE, IgD) and subclasses (IgG1, IgG2, IgG3, IgG4, IgA1 and IgA2) by indirect ELISA would required subclass-specific secondary antibodies and nine independent measurements for each patient sample. As a result, common serological tests do not evaluate the isotype- and subclass-specific humoral immune response. The identity and circulating levels of immunoglobulin isotypes and subclasses, however, vary due to time after exposure (early response IgM antibodies), antigen identity (IgG1 and IgG3 for peptide antigens and IgG2 for polysaccharides), route of infection (respiratory, urinary, or topical), cell-mediated immunity (IgM class switching to either IgG1/IgG3 or IgG2/IgG4 induced by type 1 or type 2 helper T cells, respectively), subclass stability (reduced half-life of IgG3 due to a longer hinge region and faster proteolysis), affinity (lower-affinity antibodies are not detected), and different effector functions (antibody-dependent cell-mediated cytotoxicity of IgG1>IgG3, antibody-dependent cell-mediated phagocytosis of IgG1, IgG2 and IgG3, or complement-mediated cytotoxicity of IgG1 and IgG3)^13,14^. Differential quantification of a full set of immunoglobulin isotypes and subclasses may provide additional prognostic information on disease severity and complement existing serological tests.

In future, our IP-targeted proteomics assays could be utilized widely to improve serological testing for numerous infectious diseases, such as HIV, hepatitis and influenza^67^, and evaluate quality and efficiency of vaccines^68^. With its nearly absolute analytical selectivity, mass spectrometry assays could serve as independent tools to evaluate diagnostic specificity of the existing serological tests, and select combinations of antigens and antibody subclasses which provide the highest diagnostic specificity. Increased diagnostic specificity (low false-positive rates) and standardization of serology diagnostics will enable evidence-based screening of broader populations for the acquired immunity, thus having a tremendous impact on management of infectious diseases. IP-SRM assays targeting novel antigens or mutated epitopes can be developed and implemented within weeks and are justified as tools for the rapid response to the emerging pandemics. Investigation of mutated neutralizing epitopes for their ability to bind antibodies will support development of vaccines effective against the emerging mutants. Further developments of serological diagnostics by IP-targeted proteomics assays could facilitate selection of antibodies with a desired subclass, affinity^69-71^, neutralization potential, mutation binding, and eventually provide approches for selection and sequencing of antibodies with the desired characteristics directly from the patient’s blood, thus paving the way for rapid development of the next-generation therapeutic antibodies.

## EXPERIMENTAL SECTION

### Chemicals, reagents and clinical samples

Dithiothreitol, iodoacetamide, trifluoracetic acid (TFA) and L-methionine were obtained from Thermo Fisher Scientific (Burlington, ON, Canada). Mass spectrometry-grade acetonitrile (ACN) and water were purchased from Fisher Scientific (Fair Lawn, NJ). Formic acid (FA) was obtained from Sigma-Aldrich (Oakville, ON). Synthetic stable isotope-labeled peptides SpikeTides_L and SpikeTide_TQL were provided by JPT Peptide Technologies GmbH (Germany). Recombinant SARS-CoV-2 antigens were obtained from Sino Biological (Beijing, China) and included spike glycoprotein extracellular domain (S1+S2 ECD; identical to Val16-Pro1213 of SPIKE_SARS2 (P0DTC2) with a polyhistidine tag at the C-term; 134.3 kDa; #40589-V08B1), the S1 subunit of spike glycoprotein (S1; Val16-Arg685; His-tagged, 76.5 kDa; #40591-V08H), the receptor binding domain of the spike glycoprotein (RBD; Arg319-Phe541; His-tagged, 26.54 kDa; #40592-V08H), and nucleoprotein (N; identical to Met1-Ala419 of NCAP_SARS2 (P0DTC9); His-tagged; 47.08 kDa; #40588-V08B). Anti-SPIKE_SARS2 antibodies from Sino Biological Inc included spike RBD chimeric monoclonal antibody CmAb (#40150-D001), spike S1 rabbit monoclonal RmAb R007 (#40150-R007) and spike RBD rabbit polyclonal antibody RpAb (#40592-T62). Anti-nucleoprotein antibodies from Sino Biological Inc included rabbit polyclonal antibody RpAb (# 40588-T62), rabbit monoclonal antibodies RmAb R001 (#40143-R001) and RmAb R019 (#40143-R019), and mouse monoclonal antibodies MmAb (#40143-MM05). Secondary antibodies from Invitrogen included horseradish peroxidase-conjugated polyclonal goat-anti-human IgG Fcγ (#A18817) and goat-anti-human IgG/IgM/IgA H+L (#A18847). A detailed list of antibodies is presented in **Table S1**. Seven SARS-CoV-2 negative (collected before November 2019; #ISERS2ML) and three SARS-CoV-2 positive (diagnosed by RT-PCR; #ISERSCOV2P100UL) single donor human serum samples (**Table S2**) were obtained from Innovative Research (Novi, MI, USA). The study was approved by the University of Alberta (ethics approval #Pro00104098).

### Immunoprecipitation (IP) and reversed IP

Four anti-NCAP_SARS2 and three anti-SPIKE_SARS2 antibodies were used for immunoprecipitation of recombinant NCAP_SARS2 and SPIKE_SARS2 (monomeric S1+S2 ECD; 134.3 kDa; #40589-V08B1) proteins spiked into human serum, respectively. Antibodies were diluted in phosphate-buffered saline (PBS, pH 7.4), coated onto a high-binding 96-well microplates (Greiner Bio-One) at 500 ng/well in 100 μl, and incubated overnight at RT. After washing 3 times with 200 μl wash buffer (0.1% Tween 20 in PBS), the plate was blocked for 1 h with 200 μl of blocking buffer (2% bovine serum albumin in wash buffer). The washing step was repeated. Human serum (50 μl) with spiked-in recombinant proteins was diluted to 100 μl with dilution buffer (0.1% BSA in wash buffer, 0.2 μm filtered) and added to the plates. Following 2 h incubation with continuous shaking, the plate was washed 3 times with wash buffer and 3 times with 50 mM NaHCO_3_. To measure anti-SARS-CoV-2 antibodies, four recombinant proteins (S1+S2 ECD, S1, RBD, and N) were coated overnight (500 ng per well). Positive and negative human serum samples (4 μl) were diluted 25-fold with dilution buffer, incubated (100 μl per well) for 2 h at RT, and were washed 3 times with wash buffer and 3 times with 50 mM NaHCO_3_.

### Proteomic Sample Preparation

Enriched proteins or antibodies were reduced with 10 mM dithiothreitol at 70°C for 15 min, and disulfide bonds were alkylated with 20 mM iodoacetamide at room temperature (RT) in the dark for 45 min. Heavy isotope-labeled SpikeTides_TQL (100 *f*mol per digest; **Table S3**) were used for quantification of recombinant proteins, and were spiked into each sample before digestion (0.25 ng trypsin per well). SpikeTides_L peptides (**Table S4**) were used for quantification of antibody subclasses and isotypes, and were spiked after digestion (100 *f*mol per digest). Proteins were digested overnight at 37 °C using dimethylated SOLu-trypsin (Sigma-Aldrich; 1:20 trypsin:protein). Digestion was stopped with TFA (1%), and 1 μL of 0.4 M L-methionine was added to prevent methionine oxidation during sample storage. OMIX C18 tips (10 μL; Agilent Technologies) were used for desalting and microextraction of tryptic peptides, which were then eluted with 3 μl of 65% ACN and diluted with 36 μl of 0.1% FA. Each digest was analyzed in duplicates (10 μl injections).

### Liquid Chromatography and Shotgun Mass Spectrometry Analysis

The best proteotypic peptides for S1+S2 ECD and N recombinant proteins were identified by shotgun mass spectrometry using Orbitrap Elite™ Hybrid Ion Trap-Orbitrap mass spectrometer (Thermo Scientific) coupled to EASY-nLC II (Thermo Scientific). Peptides were separated at 300 nL/min with a 2-hour gradient: 5% B for 5 min, 5-35% B for 95 min, 35-65% B for 10 min, 65-100% B for 1 min and 100% B for 9 min. MS1 scans (400-1250 m/z) were performed at 60 K resolution in the profile mode, followed by top 20 ion trap centroid MS/MS, acquired at 33% normalized collision energy. FTMS ion count was set to 1×10^6^ with an injection time of 200 ms, while MS/MS scans were set to 9,000 counts and 100 ms injection time. MS/MS acquisition settings included 500 minimum signal threshold, 2.0 m/z isolation width, 10 ms activation time, and 60 s dynamic exclusion. Monoisotopic precursor selection was enabled, +1 and unknown charge states were rejected. Instrument parameters included 230°C capillary temperature and 2.0 kV spray voltage.

### Selection of Proteotypic Peptides and Development of Targeted SRM and PRM Assays

The top 2 peptides with the highest MS1 intensities were selected for S1+S2 ECD and N recombinant proteins. To facilitate absolute quantification by PRM assay using Q-Exactive, heavy isotope labelled SpikeTides_TQL peptides were used as internal standards. To select the best proteotypic peptides for quantification of human antibodies, we used our previous shotgun mass spectrometry data. Previous literature and the Peptide Atlas, protein BLAST, neXtProt and gnomAD databases were used to confirm specificity of proteotypic peptides for the differential quantification of antibody isotypes (IgG, IgM, and IgA) and subclasses (IgG1, IgG2, IgG3, IgG4, IgA1, and IgA2), and to exclude peptides with posttranslational modifications, allotype variants and high-frequency single aminoacid variants. Heavy isotope-labeled SpikeTides_L peptides were obtained and used as internal standards for SRM assay development (**Table S4**). Heavy and light peptide pairs (10 transitions per peptide; 5 ms scan time) were initially monitored with an unscheduled SRM assay. Following deletion of low-intensity and high-interference transitions, 3 transitions for each precursor ion were scheduled within 150 s intervals (**Tables S5** and **S6**).

### Liquid chromatography and SRM/PRM parameters

Q-Exactive coupled to EASY-nLC 1000 (Thermo Scientific) was used for PRM assays. Acclaim PepMap 100 nanoViper C18 column (Thermo Scientific, 100 µm ID×2 cm, 5 μm, 100 Å) was used as a pre-column for sample loading, while the EASY-Spray C18 column (Thermo Scientific, 75 μm ID×15 cm, 3 μm, 5 μm) was used as an analytical column. An 18-min gradient (400 nL/min) started with 0% buffer B and ramped to 50% buffer B over 15 min, followed by an increase to 100% buffer B within 1 min, and continued for 2 min. PRM scans were performed at 17.5 K resolution with 27% normalized collision energy. AGC (Automatic Gain Control) target value was set to 3×10^6^ with a maximum injection time of 100 ms and an isolation width of 2.0 m/z. The quadrupole ion-trap mass spectrometer (AB SCIEX QTRAP 5500) coupled to EASY-nLC II via a NanoSpray III ion source (AB SCIEX) was used for SRM assays. The tryptic peptides were loaded at 5 μL/min onto a C18 trap column (Thermo Scientific, 100 µm ID×2 cm, 5 μm, 120 Å). Peptides were separated with PicoFrit columns (New Objective, 15 cm×75 μm ID, 8 μm tip, PepMap C18, 3 μm, 100 Å) and 18 min gradients (300 nL/min). The gradient started with 15% buffer B and ramped to 65% buffer B over 15 min, followed by an increase to 100% buffer B within 1 min, and continued for 2 min. QTRAP 5500 parameters were: 2300 V ionspray; 75 °C source temperature; 2.0 arbitrary units for gas 1 (N_2_), 0 arbitrary units for gas 2; 25 arbitrary units for curtain gas (N_2_); and 100 V declustering potential. BSA (200 *f*mol) was analyzed every 6 runs to assess nanoLC-MS performance.

### Mass spectrometry data analysis

Raw files of SRM and PRM experiments were analyzed using Skyline Targeted Proteomics Environment v20.1.0.76 (MacCoss Lab). Peak boundaries were adjusted manually, and the integrated areas of all transitions for each peptide were extracted. Light-to-heavy peak area ratios were used for accurate relative or absolute quantification of endogenous peptides. Shotgun MS data were search using MaxQuant software (v1.6.3.4) and a custom Fasta database with 29 SARS-CoV-2 proteins (NCBI Reference Sequences) and some human proteins (60 entries in total). Search parameters included: trypsin enzyme specificity, 2 missed cleavages, 7 aa minimum peptide length, top 8 MS/MS peaks per 100 Da, 20 ppm MS1 and 0.5 Da MS/MS tolerance. Variable modifications included methionine oxidation, N-terminal acetylation and deamidation (N). False-discovery rate (FDR) was set to 1% at both protein and peptide levels. The mass spectrometry proteomics data have been deposited to the ProteomeXchange Consortium via PRIDE^19^ with the dataset identifier PXD028560. Raw SRM and PRM data have been deposited to Peptide Atlas with the identifier PASS01699 (www.peptideatlas.org/PASS/PASS01699).

### ELISA

Immunoassays were developed as previously described^20^. Four proteins (S1+S2 ECD, S1, RBD, and N) were coated onto the plate overnight (300 ng in 100 μl PBS per well). The plate was washed 6 times with 250 μl wash buffer (0.1% Tween 20 in PBS), followed by 1 h blocking with 250 μl of blocking buffer (6% BSA in wash buffer). After washing, 3 positive and 7 negative human serum samples was diluted 15,000-fold with a dilution buffer (0.1% BSA in wash buffer). Samples (100 μl per well) were incubated for 2 h at RT with continuous shaking. HRP-conjugated secondary antibodies (goat-anti-human IgG and goat-anti-human IgG/IgM/IgA; 20 ng in 100 μl per well diluted in 0.5% BSA in wash buffer) were incubated for 1 h. Following final washing, 100 μl of tetramethylbenzidine (TMB) solution was added to each well. After 10 min incubation, the reaction was stopped with 50 μl of 2 M HCL. Absorbance at 450 nm was measured using FilterMax F5 multi-mode microplate reader (Molecular Devices) with 450NMBW80 absorbance filter. To measure recombinant S1 SPIKE_SARS2 and NCAP_SARS2 proteins spiked into human serum, four anti-NCAP_SARS2 and three anti-SPIKE_SARS2 antibodies (300 ng per well) were coated onto the plate. To generate standard curves, recombinant antigens (62.5 to 10,000 pg/ml) were spiked into human serum. Seven different anti-SARS2 antibodies were biotinylated in-house and used as secondary antibodies (40 ng in 100 μl per well). Following 1 h incubation, streptavidin-conjugated horseradish peroxidase (1:1000) was added and incubated for 20 min. Reaction was stopped with 50 μl of 2 M HCL, and absorbance at 450 nm was measured.

## Data Availability

All data produced in the present study are available upon reasonable request to the authors

## Acknowledgements

We thank Prof. Xingfang Li for the access to QTRAP 5500 mass spectrometer.

## Data Availability

Raw shotgun MS data have been deposited to ProteomeXchange Consortium via PRIDE (www.ebi.ac.uk/pride/archive/login) with the dataset identifier PXD028560. Raw SRM and PRM data, as well as processed Skyline files, were deposited to Peptide Atlas with the dataset identifier PASS01699.

## Funding

This work was supported by the Canadian Institutes of Health Research COVID-19 Immunity Task Force grant (#VR2-173207) and Alberta Innovates National Partnered R&I Initiatives (#202100452) to A.P.D. Z. F. acknowledges the support from the National Natural Science Foundation of China (21806018) and Globalink Early Career Fellowship (Mitacs, Canada and China Scholarship Council, China; 201806065018).

## Author contributions

A.P.D. designed the research project. Z.F. and Y.R. performed all experiments. A.P.D., Z.F. and Y.R. analyzed data. A.P.D., Z.F. and Y.R. wrote manuscript, and all authors contributed to revisions.

## Conflict of interests

The authors declare no potential conflicts of interest.

## Supplemental materials

This article contains supplemental materials.

## Notes

### Competing Interest Statement

The authors have declared no competing interest.

### Funding Statement

This study was funded by the Canadian Institutes of Health Research COVID-19 Immunity Task Force grant (#VR2-173207) and Alberta Innovates National Partnered R&I Initiatives (#202100452) to A.P.D. Z. F. acknowledges the support from the National Natural Science Foundation of China (21806018) and Globalink Early Career Fellowship (Mitacs, Canada and China Scholarship Council, China; 201806065018).

### Author Declarations

Ethics committee of University of Alberta gave ethical approval for this work (#Pro00104098).

## REFERENCES

1. Feng, W.; Newbigging, A. M.; Le, C.; Pang, B.; Peng, H.; Cao, Y.; Wu, J.; Abbas, G., et al., Molecular Diagnosis of COVID-19: Challenges and Research Needs. Anal Chem 2020, 92, 10196–10209.

2. Kanji, J. N.; Zelyas, N.; MacDonald, C.; Pabbaraju, K.; Khan, M. N.; Prasad, A.; Hu, J.; Diggle, M., et al., False negative rate of COVID-19 PCR testing: a discordant testing analysis. Virol J 2021, 18, 13.

3. Rais, Y.; Fu, Z.; Drabovich, A. P., Mass spectrometry-based proteomics in basic and translational research of SARS-CoV-2 coronavirus and its emerging mutants. Clin Proteomics 2021, 18, 19.

4. Liu, G.; Rusling, J. F., COVID-19 Antibody Tests and Their Limitations. ACS sensors 2021, 6, 593–612.

5. Pokhrel, P.; Hu, C.; Mao, H., Detecting the Coronavirus (COVID-19). ACS sensors 2020, 5, 2283–2296.

6. Wee, S.; Alli-Shaik, A.; Kek, R.; Swa, H. L. F.; Tien, W. P.; Lim, V. W.; Leo, Y. S.; Ng, L. C., et al., Multiplex targeted mass spectrometry assay for one-shot flavivirus diagnosis. Proc Natl Acad Sci U S A 2019, 116, 6754–6759.

7. Parker, J.; Carrasco, A. F.; Chen, J., BioRad BioPlex® HIV Ag-Ab assay: Incidence of false positivity in a low-prevalence population and its effects on the current HIV testing algorithm. J Clin Virol 2019, 116, 1–3.

8. Eshetu, A.; Hauser, A.; Schmidt, D.; Bartmeyer, B.; Bremer, V.; Obermeier, M.; Ehret, R.; Volkwein, A., et al., Comparison of two immunoassays for concurrent detection of HCV antigen and antibodies among HIV/HCV co-infected patients in dried serum/plasma spots. J Virol Methods 2020, 279, 113839.

9. Hwang, J. P.; Feld, J. J.; Hammond, S. P.; Wang, S. H.; Alston-Johnson, D. E.; Cryer, D. R.; Hershman, D. L.; Loehrer, A. P., et al., Hepatitis B Virus Screening and Management for Patients With Cancer Prior to Therapy: ASCO Provisional Clinical Opinion Update. J Clin Oncol 2020, 38, 3698–3715.

10. Chen, Z.; Zhang, Z.; Zhai, X.; Li, Y.; Lin, L.; Zhao, H.; Bian, L.; Li, P., et al., Rapid and Sensitive Detection of anti-SARS-CoV-2 IgG, Using Lanthanide-Doped Nanoparticles-Based Lateral Flow Immunoassay. Anal Chem 2020, 92, 7226–7231.

11. Faustini, S. E.; Jossi, S. E.; Perez-Toledo, M.; Shields, A. M.; Allen, J. D.; Watanabe, Y.; Newby, M. L.; Cook, A., et al., Development of a high-sensitivity ELISA detecting IgG, IgA and IgM antibodies to the SARS-CoV-2 spike glycoprotein in serum and saliva. Immunology 2021, 164, 135–147.

12. Liu, G.; Rusling, J. F., COVID-19 Antibody Tests and Their Limitations. ACS Sens 2021, 6, 593–612.

13. Xu, Z.; Zan, H.; Pone, E. J.; Mai, T.; Casali, P., Immunoglobulin class-switch DNA recombination: induction, targeting and beyond. Nat Rev Immunol 2012, 12, 517–531.

14. Vidarsson, G.; Dekkers, G.; Rispens, T., IgG subclasses and allotypes: from structure to effector functions. Front Immunol 2014, 5, 520.

15. Zecha, J.; Lee, C. Y.; Bayer, F. P.; Meng, C.; Grass, V.; Zerweck, J.; Schnatbaum, K.; Michler, T., et al., Data, Reagents, Assays and Merits of Proteomics for SARS-CoV-2 Research and Testing. Mol Cell Proteomics 2020, 19, 1503–1522.

16. Cardozo, K. H. M.; Lebkuchen, A.; Okai, G. G.; Schuch, R. A.; Viana, L. G.; Olive, A. N.; Lazari, C. D. S.; Fraga, A. M., et al., Establishing a mass spectrometry-based system for rapid detection of SARS-CoV-2 in large clinical sample cohorts. Nat Commun 2020, 11, 6201.

17. Cazares, L. H.; Chaerkady, R.; Samuel Weng, S. H.; Boo, C. C.; Cimbro, R.; Hsu, H. E.; Rajan, S.; Dall’Acqua, W., et al., Development of a Parallel Reaction Monitoring Mass Spectrometry Assay for the Detection of SARS-CoV-2 Spike Glycoprotein and Nucleoprotein. Anal Chem 2020, 92, 13813–13821.

18. Ihling, C.; Tanzler, D.; Hagemann, S.; Kehlen, A.; Huttelmaier, S.; Arlt, C.; Sinz, A., Mass Spectrometric Identification of SARS-CoV-2 Proteins from Gargle Solution Samples of COVID-19 Patients. J Proteome Res 2020, 19, 4389–4392.

19. Perez-Riverol, Y.; Csordas, A.; Bai, J.; Bernal-Llinares, M.; Hewapathirana, S.; Kundu, D. J.; Inuganti, A.; Griss, J., et al., The PRIDE database and related tools and resources in 2019: improving support for quantification data. Nucleic Acids Res 2019, 47, D442–D450.

20. Isho, B.; Abe, K. T.; Zuo, M.; Jamal, A. J.; Rathod, B.; Wang, J. H.; Li, Z.; Chao, G., et al., Persistence of serum and saliva antibody responses to SARS-CoV-2 spike antigens in COVID-19 patients. Sci Immunol 2020, 5.

21. Fu, Z.; Drabovich, A. P., LC-MS/MS analysis of recombinant Spike and Nucleocapsid proteins [Data set]. Zenodo 2020, https://doi.org/10.5281/zenodo.3743880.

22. Grossegesse, M.; Hartkopf, F.; Nitsche, A.; Schaade, L.; Doellinger, J.; Muth, T., Perspective on Proteomics for Virus Detection in Clinical Samples. J Proteome Res 2020, 19, 4380–4388.

23. Renuse, S.; Vanderboom, P. M.; Maus, A. D.; Kemp, J. V.; Gurtner, K. M.; Madugundu, A. K.; Chavan, S.; Peterson, J. A., et al., A mass spectrometry-based targeted assay for detection of SARS-CoV-2 antigen from clinical specimens. EBioMedicine 2021, 69, 103465.

24. Shan, D.; Johnson, J. M.; Fernandes, S. C.; Suib, H.; Hwang, S.; Wuelfing, D.; Mendes, M.; Holdridge, M., et al., N-protein presents early in blood, dried blood and saliva during asymptomatic and symptomatic SARS-CoV-2 infection. Nat Commun 2021, 12, 1931.

25. Schiza, C.; Korbakis, D.; Jarvi, K.; Diamandis, E. P.; Drabovich, A. P., Identification of TEX101-associated proteins through proteomic measurement of human spermatozoa homozygous for the missense variant rs35033974. Mol Cell Proteomics 2019, 18, 338.

26. Kusebauch, U.; Campbell, D. S.; Deutsch, E. W.; Chu, C. S.; Spicer, D. A.; Brusniak, M. Y.; Slagel, J.; Sun, Z., et al., Human SRMAtlas: A Resource of Targeted Assays to Quantify the Complete Human Proteome. Cell 2016, 166, 766–778.

27. Ladwig, P. M.; Barnidge, D. R.; Snyder, M. R.; Katzmann, J. A.; Murray, D. L., Quantification of serum IgG subclasses by use of subclass-specific tryptic peptides and liquid chromatography--tandem mass spectrometry. Clin Chem 2014, 60, 1080–1088.

28. Remily-Wood, E. R.; Benson, K.; Baz, R. C.; Chen, Y. A.; Hussein, M.; Hartley-Brown, M. A.; Sprung, R. W.; Perez, B., et al., Quantification of peptides from immunoglobulin constant and variable regions by LC-MRM MS for assessment of multiple myeloma patients. Proteomics Clin Appl 2014, 8, 783–795.

29. Karczewski, K. J.; Francioli, L. C.; Tiao, G.; Cummings, B. B.; Alfoldi, J.; Wang, Q.; Collins, R. L.; Laricchia, K. M., et al., The mutational constraint spectrum quantified from variation in 141,456 humans. Nature 2020, 581, 434–443.

30. Gaudet, P.; Michel, P.-A.; Zahn-Zabal, M.; Britan, A.; Cusin, I.; Domagalski, M.; Duek, P. D.; Gateau, A., et al., The neXtProt knowledgebase on human proteins: 2017 update. Nucleic acids research 2017, 45, D177–D182.

31. Premkumar, L.; Segovia-Chumbez, B.; Jadi, R.; Martinez, D. R.; Raut, R.; Markmann, A.; Cornaby, C.; Bartelt, L., et al., The receptor binding domain of the viral spike protein is an immunodominant and highly specific target of antibodies in SARS-CoV-2 patients. Sci Immunol 2020, 5.

32. Drabovich, A. P.; Okhonin, V.; Berezovski, M.; Krylov, S. N., Smart aptamers facilitate multi-probe affinity analysis of proteins with ultra-wide dynamic range of measured concentrations. J Am Chem Soc 2007, 129, 7260–7261.

33. Cassidy, J. T.; Nordby, G. L., Human serum immunoglobulin concentrations: prevalence of immunoglobulin deficiencies. J Allergy Clin Immunol 1975, 55, 35–48.

34. French, M. A.; Harrison, G., Systemic antibody deficiency in patients without serum immunoglobulin deficiency or with selective IgA deficiency. Clin Exp Immunol 1984, 56, 18–22.

35. Puissant-Lubrano, B.; Peres, M.; Apoil, P. A.; Congy-Jolivet, N.; Roubinet, F.; Blancher, A., Immunoglobulin IgA, IgD, IgG, IgM and IgG subclass reference values in adults. Clin Chem Lab Med 2015, 53, e359–361.

36. Sethuraman, N.; Jeremiah, S. S.; Ryo, A., Interpreting Diagnostic Tests for SARS-CoV-2. JAMA 2020, 323, 2249–2251.

37. Brownstein, N. C.; Chen, Y. A., Predictive values, uncertainty, and interpretation of serology tests for the novel coronavirus. Sci Rep 2021, 11, 5491.

38. Bar-On, Y. M.; Flamholz, A.; Phillips, R.; Milo, R., SARS-CoV-2 (COVID-19) by the numbers. Elife 2020, 9.

39. Li, Y. H.; Li, J.; Liu, X. E.; Wang, L.; Li, T.; Zhou, Y. H.; Zhuang, H., Detection of the nucleocapsid protein of severe acute respiratory syndrome coronavirus in serum: comparison with results of other viral markers. J Virol Methods 2005, 130, 45–50.

40. Drabovich, A. P.; Martinez-Morillo, E.; Diamandis, E. P., Toward an integrated pipeline for protein biomarker development. Biochim Biophys Acta 2015, 1854, 677–686.

41. Drabovich, A. P.; Pavlou, M. P.; Batruch, I.; Diamandis, E. P., Chapter 2 - Proteomic and mass spectrometry technologies for biomarker discovery. In Proteomic and Metabolomic Approaches to Biomarker Discovery, Issaq, H. J.; Veenstra, T. D., Eds., Academic Press (Elsevier): Waltham, MA, 2013, pp 17–37.

42. Drabovich, A. P.; Saraon, P.; Jarvi, K.; Diamandis, E. P., Seminal plasma as a diagnostic fluid for male reproductive system disorders. Nat Rev Urol 2014, 11, 278–288.

43. Drabovich, A. P.; Martínez-Morillo, E.; Diamandis, E. P. Protein Biomarker Discovery. In Proteomics for Biological Discovery, 2019, pp 63–88.

44. Karakosta, T. D.; Soosaipillai, A.; Diamandis, E. P.; Batruch, I.; Drabovich, A. P., Quantification of human kallikrein-related peptidases in biological fluids by multiplatform targeted mass spectrometry assays. Mol Cell Proteomics 2016, 15, 2863.

45. Drabovich, A. P.; Dimitromanolakis, A.; Saraon, P.; Soosaipillai, A.; Batruch, I.; Mullen, B.; Jarvi, K.; Diamandis, E. P., Differential diagnosis of azoospermia with proteomic biomarkers ECM1 and TEX101 quantified in seminal plasma. Sci Transl Med 2013, 5, 212ra160.

46. Drabovich, A. P.; Jarvi, K.; Diamandis, E. P., Verification of male infertility biomarkers in seminal plasma by multiplex selected reaction monitoring assay. Mol Cell Proteomics 2011, 10, M110 004127.

47. Korbakis, D.; Brinc, D.; Schiza, C.; Soosaipillai, A.; Jarvi, K.; Drabovich, A. P.; Diamandis, E. P., Immunocapture-selected reaction monitoring screening facilitates the development of ELISA for the measurement of native TEX101 in biological fluids. Mol Cell Proteomics 2015, 14, 1517–1526.

48. Drabovich, A. P.; Saraon, P.; Drabovich, M.; Karakosta, T. D.; Dimitromanolakis, A.; Hyndman, M. E.; Jarvi, K.; Diamandis, E. P., Multi-omics Biomarker Pipeline Reveals Elevated Levels of Protein-glutamine Gamma-glutamyltransferase 4 in Seminal Plasma of Prostate Cancer Patients. Mol Cell Proteomics 2019, 18, 1807–1823.

49. Schiza, C.; Korbakis, D.; Panteleli, E.; Jarvi, K.; Drabovich, A. P.; Diamandis, E. P., Discovery of a human testis-specific protein complex TEX101-DPEP3 and selection of its disrupting antibodies. Mol Cell Proteomics 2018, 17, 2480–2495.

50. Drabovich, A. P.; Pavlou, M. P.; Dimitromanolakis, A.; Diamandis, E. P., Quantitative analysis of energy metabolic pathways in MCF-7 breast cancer cells by selected reaction monitoring assay. Mol Cell Proteomics 2012, 11, 422–434.

51. Drabovich, A. P.; Pavlou, M. P.; Schiza, C.; Diamandis, E. P., Dynamics of protein expression reveals primary targets and secondary messengers of estrogen receptor alpha signaling in MCF-7 breast cancer cells. Mol Cell Proteomics 2016, 15, 2093–2107.

52. Konvalinka, A.; Zhou, J.; Dimitromanolakis, A.; Drabovich, A. P.; Fang, F.; Gurley, S.; Coffman, T.; John, R., et al., Determination of an angiotensin II-regulated proteome in primary human kidney cells by stable isotope labeling of amino acids in cell culture (SILAC). J Biol Chem 2013, 288, 24834–24847.

53. Korbakis, D.; Schiza, C.; Brinc, D.; Soosaipillai, A.; Karakosta, T. D.; Legare, C.; Sullivan, R.; Mullen, B., et al., Preclinical evaluation of a TEX101 protein ELISA test for the differential diagnosis of male infertility. BMC Med 2017, 15, 60.

54. Cho, C. K.; Drabovich, A. P.; Karagiannis, G. S.; Martinez-Morillo, E.; Dason, S.; Dimitromanolakis, A.; Diamandis, E. P., Quantitative proteomic analysis of amniocytes reveals potentially dysregulated molecular networks in Down syndrome. Clin Proteomics 2013, 10, 2.

55. Fu, Z.; Rais, Y.; Bismar, T. A.; Hyndman, M. E.; Le, X. C.; Drabovich, A. P., Mapping Isoform Abundance and Interactome of the Endogenous TMPRSS2-ERG Fusion Protein by Orthogonal Immunoprecipitation-Mass Spectrometry Assays. Mol Cell Proteomics 2021, 20, 100075.

56. Begcevic, I.; Brinc, D.; Drabovich, A. P.; Batruch, I.; Diamandis, E. P., Identification of brain-enriched proteins in the cerebrospinal fluid proteome by LC-MS/MS profiling and mining of the Human Protein Atlas. Clin Proteomics 2016, 13, 11.

57. Begcevic, I.; Brinc, D.; Dukic, L.; Simundic, A. M.; Zavoreo, I.; Basic Kes, V.; Martinez-Morillo, E.; Batruch, I., et al., Targeted mass spectrometry-based assays for relative quantification of 30 brain-related proteins and their clinical applications. J Proteome Res 2018, 17, 2282–2292.

58. Martinez-Morillo, E.; Cho, C. K.; Drabovich, A. P.; Shaw, J. L.; Soosaipillai, A.; Diamandis, E. P., Development of a multiplex selected reaction monitoring assay for quantification of biochemical markers of down syndrome in amniotic fluid samples. J Proteome Res 2012, 11, 3880–3887.

59. Martinez-Morillo, E.; Nielsen, H. M.; Batruch, I.; Drabovich, A. P.; Begcevic, I.; Lopez, M. F.; Minthon, L.; Bu, G., et al., Assessment of peptide chemical modifications on the development of an accurate and precise multiplex selected reaction monitoring assay for apolipoprotein e isoforms. J Proteome Res 2014, 13, 1077–1087.

60. Cho, C. K.; Drabovich, A. P.; Batruch, I.; Diamandis, E. P., Verification of a biomarker discovery approach for detection of Down syndrome in amniotic fluid via multiplex selected reaction monitoring (SRM) assay. J Proteomics 2011, 74, 2052–2059.

61. Konvalinka, A.; Batruch, I.; Tokar, T.; Dimitromanolakis, A.; Reid, S.; Song, X.; Pei, Y.; Drabovich, A. P., et al., Quantification of angiotensin II-regulated proteins in urine of patients with polycystic and other chronic kidney diseases by selected reaction monitoring. Clin Proteomics 2016, 13, 16.

62. Drabovich, A. P.; Diamandis, E. P., Combinatorial peptide libraries facilitate development of multiple reaction monitoring assays for low-abundance proteins. J Proteome Res 2010, 9, 1236–1245.

63. Frosner, G. G.; Schomerus, H.; Wiedmann, K. H.; Zachoval, R.; Bayerl, B.; Backer, U.; Gathof, G. A.; Sugg, U., Diagnostic significance of quantitative determination of hepatitis B surface antigen in acute and chronic hepatitis B infection. Eur J Clin Microbiol 1982, 1, 52–58.

64. Che, X. Y.; Qiu, L. W.; Pan, Y. X.; Wen, K.; Hao, W.; Zhang, L. Y.; Wang, Y. D.; Liao, Z. Y., et al., Sensitive and specific monoclonal antibody-based capture enzyme immunoassay for detection of nucleocapsid antigen in sera from patients with severe acute respiratory syndrome. J Clin Microbiol 2004, 42, 2629–2635.

65. Moore, J. P.; Cao, Y.; Ho, D. D.; Koup, R. A., Development of the anti-gp120 antibody response during seroconversion to human immunodeficiency virus type 1. J Virol 1994, 68, 5142–5155.

66. Norman, M.; Gilboa, T.; Ogata, A. F.; Maley, A. M.; Cohen, L.; Busch, E. L.; Lazarovits, R.; Mao, C. P., et al., Ultrasensitive high-resolution profiling of early seroconversion in patients with COVID-19. Nat Biomed Eng 2020, 4, 1180–1187.

67. Williams, T. L.; Luna, L.; Guo, Z.; Cox, N. J.; Pirkle, J. L.; Donis, R. O.; Barr, J. R., Quantification of influenza virus hemagglutinins in complex mixtures using isotope dilution tandem mass spectrometry. Vaccine 2008, 26, 2510–2520.

68. Tarasov, M.; Shanko, A.; Kordyukova, L.; Katlinski, A., Characterization of Inactivated Influenza Vaccines Used in the Russian National Immunization Program. Vaccines (Basel) 2020, 8.

69. Drabovich, A.; Berezovski, M.; Krylov, S. N., Selection of smart aptamers by equilibrium capillary electrophoresis of equilibrium mixtures (ECEEM). J Am Chem Soc 2005, 127, 11224–11225.

70. Drabovich, A. P.; Berezovski, M.; Okhonin, V.; Krylov, S. N., Selection of smart aptamers by methods of kinetic capillary electrophoresis. Anal Chem 2006, 78, 3171–3178.

71. Drabovich, A. P.; Berezovski, M. V.; Musheev, M. U.; Krylov, S. N., Selection of smart small-molecule ligands: the proof of principle. Anal Chem 2009, 81, 490–494.

